# Detection of SARS-CoV-2 in Different Human Biofluids Using the Loop-Mediated Isothermal Amplification Assay: A Prospective Diagnostic Study in Fortaleza, Brazil

**DOI:** 10.1101/2022.01.23.22269711

**Authors:** Marco Clementino, Karene Ferreira Cavalcante, Vania Angelica Feitosa Viana, Dayara de Oliveira Silva, Caroline Rebouças Damasceno, Jessica Fernandes de Souza, Rafhaella Nogueira Della Guardia Gondim, Daniel Macedo de Melo Jorge, Lyvia Maria Vasconcelos Carneiro Magalhães, Érico Antônio Gomes de Arruda, Roberto da Justa Pires Neto, Melissa Soares Medeiros, Armênio Aguiar dos Santos, Pedro Jorge Caldas Magalhães, Liana Perdigão Mello, Eurico Arruda, Aldo Ângelo Moreira Lima, Alexandre Havt

## Abstract

We adopted the reverse transcriptase - loop mediated isothermal amplification (RT-LAMP) to detect SARS-Cov-2 in patient samples. Two primer sets for genes *N* and *Orf1ab* were designed to detect SARS-CoV-2, and one primer set was designed to detect the human gene *Actin*. We collected prospective 138 nasopharyngeal swabs, 70 oropharyngeal swabs, 69 saliva, and 68 mouth saline wash samples from patients suspected to have severe acute respiratory syndrome (SARS) caused by SARS-CoV-2 to test the RT-LAMP in comparison with the golden standard technique RT-qPCR. Accuracy of diagnosis using both primers, N5 and Orf9, was evaluated. Sensitivity and specificity for diagnosis was 96% (95% CI 87-99) and 85% (95% CI 76-91) in 138 samples, respectively. Accurate diagnosis results were obtained only in nasopharyngeal swab processed via extraction kit. Accurate and rapid diagnosis could aid COVID-19 pandemic management by identifying, isolating, and treating patients rapidly.

**Highlights:**

- New nucleic acid amplification test for the diagnosis of SARS-CoV-2 using the RT-LAMP
- N5 primer set showed mutations in strains of interest, such as the gamma strain (P.1) of SARS-CoV-2
- When evaluated in combination N5 and Orf9 primer sets maintained high sensitivity and specificity

## Introduction

The American CDC and WHO recommend quantitative polymerase chain reaction (qPCR) as the gold standard nucleic acid amplification test (NAAT) for the molecular diagnosis of infectious diseases. However, alternative methods that can circumvent the time-consuming limitations and peculiar demands of laboratory infrastructure for qPCR should be considered, especially in developing countries. Ceará is one of the Brazilian states most affected by the COVID-19 pandemic, with an incidence of 6,434 per 100,000 inhabitants based on national official data in October 2021. The government institution responsible for the diagnosis of COVID-19 in Ceará is the Central Laboratory (LACEN-CE), which uses a qPCR routine that requires an average of 48–72 h to confirm the presence of SARS-CoV-2 in patient samples.

In this study, we partnered with LACEN-CE to develop a diagnostic test for SARS-CoV-2 using the reverse transcription loop-mediated isothermal amplification (RT-LAMP) technique [1]. We propose that RT-LAMP would be faster, more accurate, and more reliable technique to detect SARS-CoV-2 in patient samples compared to conventional tests. LAMP is a single-step reaction that amplifies DNA or RNA using a single temperature of 65 °C, and a total time of execution of 40–60 min. At the end of the amplification phase, the increased turbidity of the reaction would allow us to observe the result even with the naked eye.

Several scientific articles have already been published using the LAMP technique for the molecular diagnosis of pathogens [2-8]. Since the recent SARS-CoV-2 pandemic, several studies have been published worldwide on the methods of diagnosing COVID-19 using the RT-LAMP technique [9-13]. Such an accurate and rapid diagnosis could aid in COVID-19 pandemic management by identifying, isolating, and treating patients in a timely manner and could reduce LANCEN-CE time for diagnosis by half, removing the software analysis requirements.

## Materials and Methods

### Primer design

Primers were designed for the recognition of genes *Orf1ab and N* of SARS-CoV-2 using the PrimerExplorer version 5. Specificity and limit of detection of the primers were tested using plasmid positive controls for SARS-CoV-2 genes *N* and *Orf1ab and human RNaseP* subunit 30 (*HRP30*) gene (IDT, Newark, NJ, USA) or from a human *actin* gene RT-qPCR amplicon produced with specific external LAMP primers F3 and B3. Best primers were selected for cross-specificity and limits of detection using other respiratory viruses and SARS-CoV-2 variants cultivated *in vitro*.

### Cultivation and isolation of viruses in vitro

Influenza, rhinovirus serotype 16 (RV-16)-1A and -2, human metapneumovirus (HMPV), and SARS-CoV-2 Brazil/SP-BR02/2020 virus stocks were propagated in cell cultures. Stocks were titrated by plaque assay and median tissue culture infectious dose (TCID_50_) and then aliquoted and frozen at –80 °C.

### RT-LAMP reaction

We used the WarmStart Colorimetric RT-LAMP 2X Master Mix (New England Biolabs, Ipswich, MA, USA) for the RT-LAMP reaction. The RT-LAMP protocol features a specific saline buffer, a reverse transcriptase enzyme, a Bst DNA polymerase and six specific primers (FIP, BIP, F3, B3, Loop F, and Loop B). Each set of primers were packaged initially at 10X concentration. All LAMP reactions were performed in a final volume of 25 μL. The total volume consisted of 12.5 μL of the RT-LAMP enzyme mix, 2.5 μL of the 10X concentrated primer pool, 5 μL of RNAse and DNAse free water, and 5 μL of the tested sample. The plates were sealed and heated to 65 °C for 60 min. For the commercial kit used in this study the positive samples turned yellow, while the negative samples remained pink in color.

### RT-qPCR diagnosis

The gold standard method for COVID-19 diagnosis was performed using the RT-qPCR comprising primers and probes specific for SARS-CoV-2 *N* and human genes (IDT, Newark, NJ, USA). Diagnosis performed in conjunction with a standard curve for viral load. Patient samples viral load was estimated using standard curves with efficiencies ranging from 95–105%. Samples with a viral load of ≥ 1 copy were considered positive for SARS-CoV-2 by RT-qPCR.

### Mutation rates of primers

The chosen primers set for the genes *N* and *Orf1ab* were subjected to mutation rate analysis. We selected 3844 sequences from Brazilian SARS-CoV-2 complete genome sequences from GISAID on March 18, 2021. These sequences were aligned using the Clustal W software (Ver. 1.2.4) [14] and mutation rates were calculated using the following formula: mutation rates = number of sequences with mutations/total number of sequences.

### Ethical approval

This proposal was approved by the National Ethical Review Board on 06/10/2020 with the code CAAE 33460220.7.0000.5045. Data collected from the research participants were kept confidential.

### Sample size and statistical analysis

Estimation of the required population size for this proposal was calculated with statistical power set to at least 80%, and confidence was set to be at least 95% [15, 16]. Considering the prevalence of COVID-19 as 50% positive, we estimated that 62 patients suspected of SARS-CoV-2 would be sufficient to determine the sensitivity and specificity of the test. Having a natural loss of 15% of these patients, a total of 71 patient samples were collected [17].

RT-LAMP results were classified as true positive (TP), false positive (FP), false negative (FN), or true negative (TN) based on the RT-qPCR test results using RNA extracted from nasopharyngeal samples as templates. Positive predictive values (PPV), positive negative values (PNV), sensitivity, and specificity were calculated using GraphPad Prism software (ver. 8.4.0) [18].

### Prospective study design

This study was divided into two stages to test the detection accuracy of SARS-CoV-2 in human biofluids of SARS suspected patients in Ceará, a northeastern state of Brazil.

Patients of the first stage of this study were screened for SARS symptoms in primary care units in the state of Ceará. Then, 71 SARS suspected individuals’ nasopharyngeal swabs samples were collected and aliquoted blindly by our collaborators at LACEN-CE. During the second stage of this study, patients were screened for SARS symptoms by health care employees at the São José Hospital, a public hospital in Fortaleza, Ceara, Brazil. Our second set of 71 patients’ samples suspected to have the SARS-CoV-2 infection comprised nasopharyngeal swabs, oropharyngeal swabs, saliva, and mouthwash with saline solution. Study population demographics is shown in Supplementary table 1.

Prior to diagnosis, 300 µL of the nasopharyngeal swab samples was used for RNA extraction via automatic magnetic beads RNA isolation using CHEMAGIC 360-D (PerkinElmer, Waltham, MA, USA) or MagMax viral RNA isolation kit (Applied Biosciences, Los Angeles, LA, USA). The isolated RNA was used for SARS-CoV-2 diagnosis via RT-qPCR and RT-LAMP assays. The remaining swab volume (600 μL) of nasopharyngeal swabs of stage 1 were heated at 98 °C for 30 min to isolate RNA from samples by the boiling method [19]. Afterwards, the samples were centrifuged at 5000 × g for 5 min at 4 °C. Supernatants were collected and submitted for SARS-CoV-2 diagnosis via RT-qPCR and RT-LAMP assays.

## Results

### SARS-CoV-2 diagnosis kit primer design

Five sets of primers were designed for the *N* gene of SARS-CoV-2, but only N5 reacted exclusively to the *N* gene-positive control (yellow color in **Figure 1A**). The *in vitro* limit of detection for N5 reached one copy of its positive control (IDT, Newark, NJ, USA) (**Figure 1B**). Four sets of primers were designed for the *Orf1ab* gene of SARS-CoV-2, but only Orf9 reacted exclusively to the Orf1ab positive control (IDT, Newark, NJ, USA) (yellow color in **Figure 1C**). The *in vitro* limit of detection for Orf9 was 1000 copies of its positive control (**Figure 1D**). The results showed that primers set N5 and Orf9 could be used to detect SARS-CoV-2 using sequences of SARS-CoV-2 plasmids.

**Figure 1.**
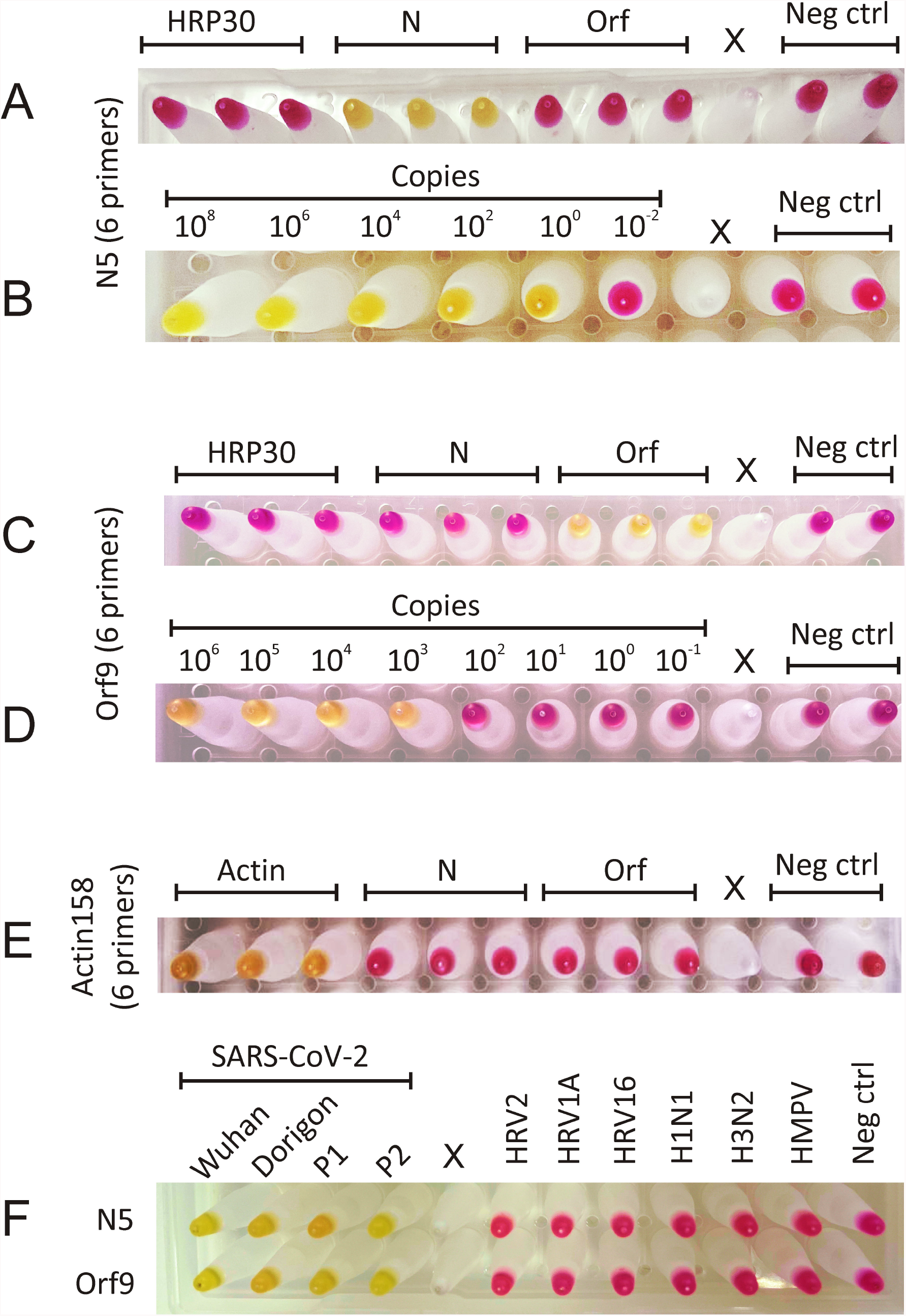
**Reaction specificity and detection limit tests. A - Representative results of the specificity of the RT-LAMP reaction using the N5 primer set and control plasmids for the human HRP30 gene, the *N* gene of SARS-Cov-2 and for the *Orf1ab* gene of SARS-CoV-2. B – Representative results of the detection limit of the RT-LAMP reaction using the N5 primer set and the control plasmid for the SARS-CoV-2 *N* gene. C - Representative results of the specificity of the RT-LAMP reaction using the primer set Orf9 and the control plasmids for the human *HRP30* gene, the SARS-Cov-2 *Orf1ab* gene and the *N* gene of SARS-CoV-2. D - Representative results of the detection limit of the RT-LAMP reaction of the Orf9 primer set and the control plasmid for the SARS-CoV-2 Orf1ab gene. E - Representative results of the specificity of RT-LAMP reaction using primer set Actin158 and positive control for human Actin gene and control plasmids for SARS-Cov-2 gene *N* and SARS-CoV-2 gene *Orf1ab*. Negative Control, Neg Ctrl; X, empty well. F – Representative results of specificity of the RT-LAMP reaction with viral samples cultured in vitro using primer sets N5 and Orf9.**

To complete the diagnostic kit, we used a primer set to detect a human gene to use as an internal control. Five sets of human actin primers were designed. The Actin158 primer set showed specificity for the positive control and were selected for our study (yellow color in **Figure 1E**).

We confirmed the reaction specificity by testing the RT-LAMP primer sets N5 and Orf9 in a reaction with other respiratory viruses obtained *in vitro*. The results showed that the new RT-LAMP primer sets developed for the detection of SARS-CoV-2 were reactive to SARS-CoV-2 isolates from different variants (Wuhan, Dorigan, P1, and P2) (yellow color in **Figure 1F**). Additionally, the new RT-LAMP primer sets did not react with other viruses tested (**Figure 1F**).

Next, we investigated primer sets mutation rates. The mutation rates of the N5 primers (F3, B3, FIP, BIP, and LF) were lower than 2%; however, the mutation rate of the N5 primer LB was 16.8% (**Figure 2A**). We observed that ∼50% of these mutations were found in clades GR and P1 (**Figure 2B** and **2C**). Alignment analysis of the Orf9 primer set showed mutation rates lower than 2% for all six primers (**Figure 2D**). Alignment of the SARS-CoV-2 genome with the N5 primer set showed a single nucleotide mutation in clades GR and P1. The mutation was characterized as a single nucleotide substitution of the nucleotide guanine for cytosine (**Figure 2E**). To test the hypothesis of higher false-negative rates of primer set N5, we tested both primer sets in an RT-LAMP reaction with serial dilutions of SARS-CoV-2 variants cultured *in vitro*. The results confirmed that primer set N5 was less reliable than Orf9, as it had a lower limit of detection (**Figure 2F and 2G**). Considering these results, we would recommend using both primer sets for the detection of SARS-CoV-2 in human samples.

**Figure 2.**
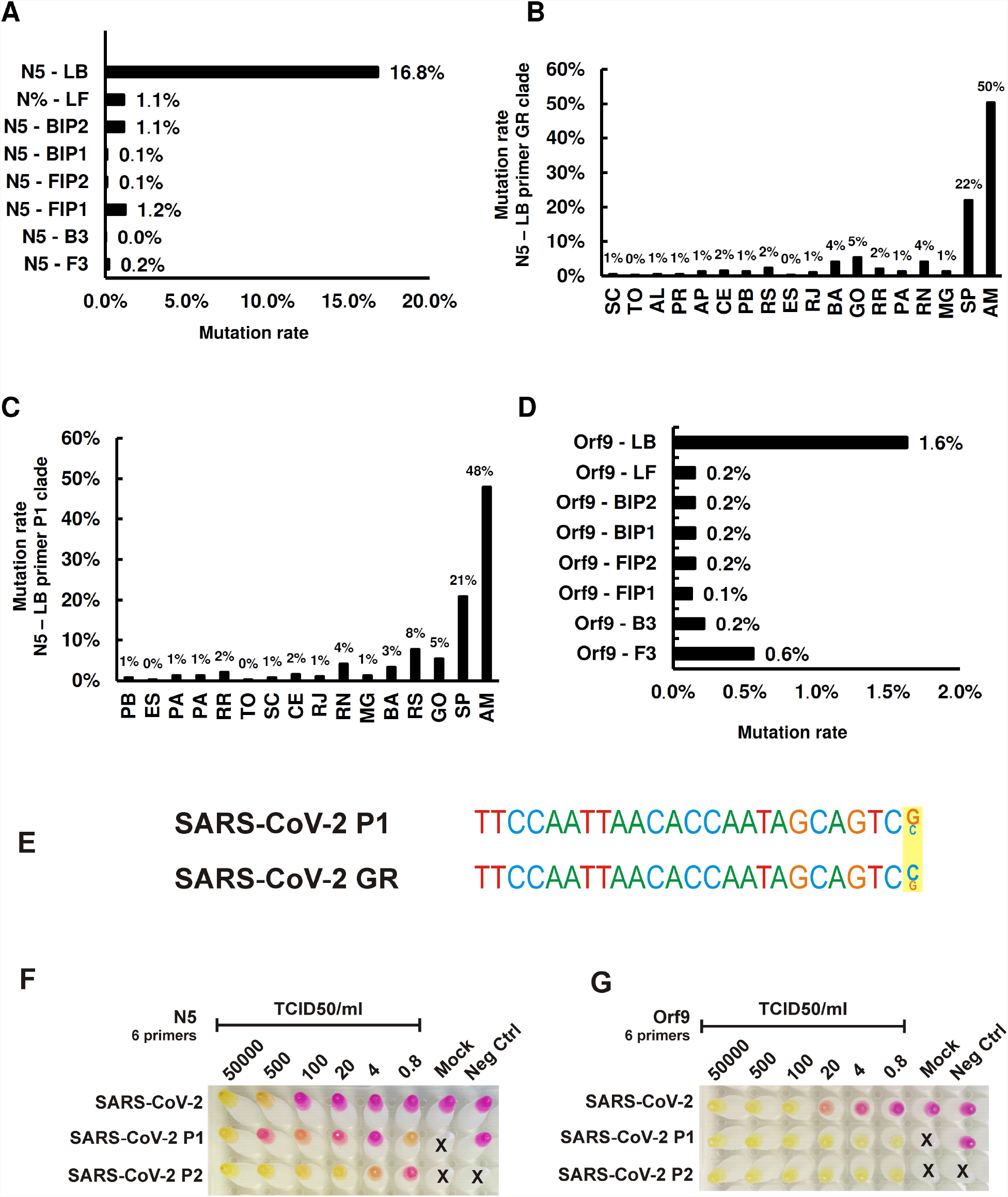
**Primer sets mutation rate. A - Mutation rate of the N5 primer set obtained from the alignment analysis of 3844 Brazilian SARS-CoV-2 sequences. B - N5-LB primer mutation rate in the GR clade by Brazilian state. C - Mutation rate of primer N5-LB in the clade P1 by Brazilian state. D - Mutation rate of the Orf9 primer set obtained from the alignment analysis of 3844 Brazilian SARS-CoV-2 sequences. E – Consensus sequence of mutations in the N5-LB sequence. F – Representative limit of detection results of the RT-LAMP reaction using the N5 primer set in samples of SARS-CoV-2, Variant P1 and variant P2 cultured in vitro. G – Representative results of the detection limit of the RT-LAMP reaction using the Orf9 primer set in samples of SARS-CoV-2, Variant P1 and Variant P2 cultured in vitro.**

### Alternative methodologies for the detection of SARS-CoV-2 via RT-qPCR and RT-LAMP

During the first stage of this study, we investigated whether different RNA extraction methods could influence the detection of SARS-CoV-2 in nasopharyngeal samples. Thus, we performed RNA isolation using an RNA isolation kit with magnetic beads and the boiling method (i.e., heat). The results showed an increase in the CT range for RT-qPCR reactions of both *Orf1ab* and *N* in samples extracted by heat, as their CT median was higher than 30. In contrast, the samples extracted using the commercial extraction kit had a CT median of approximately 22 cycles (**Figures 3A and 3B**). As expected, we also observed a significant decrease in viral load in nasopharyngeal samples extracted by heat treatment (**Figure 3C and 3D**). Overall, these results demonstrate that the boiling method did not preserve viral RNA in the nasopharyngeal samples used in this study.

**Figure 3.**
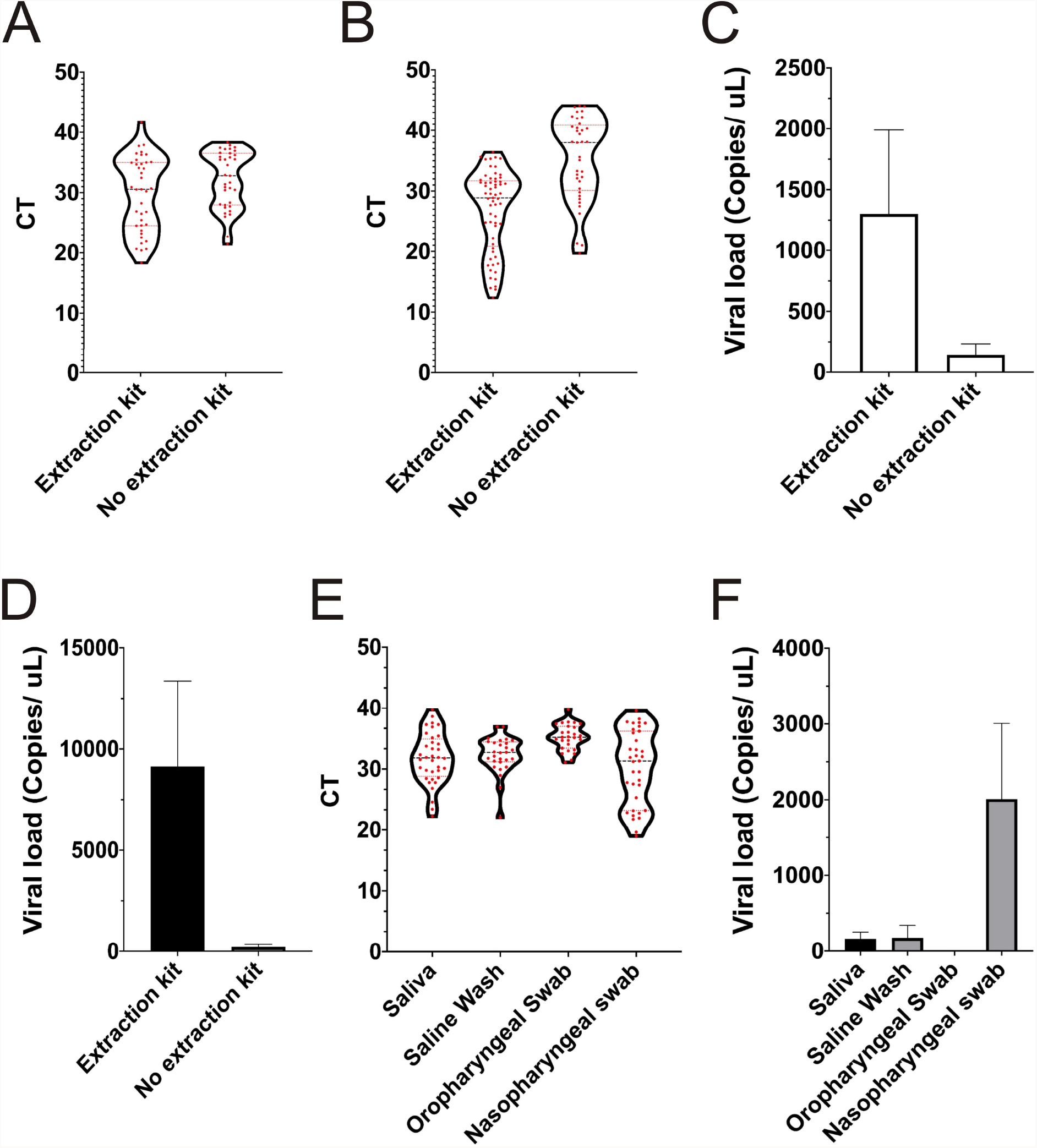
***Alternative methodologies for the detection of SARS-CoV-2 via RT-qPCR and RT-LAMP*. A – Violin plot of CT values obtained from RT-qPCR of nasopharyngeal swab samples whose RNA was extracted via RNA extraction kit or extracted via the boiling method using primers for the detection of the *Orf1ab* gene of SARS-CoV-2. B – Box plot of CT values obtained from RT-qPCR of nasopharyngeal swab samples whose RNA was extracted via RNA extraction kit or extracted via the boiling method using primers for the detection of the *N* gene of SARS-CoV-2. C – Mean of viral load obtained from RT-qPCR of nasopharyngeal swab samples whose extracted via RNA extraction kit or extracted via the boiling method using primers for the detection of the *Orf1ab* gene of SARS-CoV-2. D – Mean of viral load obtained from RT-qPCR of nasopharyngeal swab samples whose RNA was extracted via RNA extraction kit or extracted via the boiling method using primers for the detection of the *N* gene of SARS-CoV-2. E – Box plot of CT values obtained from RT-qPCR of Saliva, Saline mouth wash, Oropharyngeal swab, and nasopharyngeal swab samples whose RNA was extracted via RNA extraction kit using primers for the detection of the *Orf1ab* gene of SARS-CoV-2. F - Mean of viral load obtained from RT-qPCR of Saliva, Saline mouth wash, Oropharyngeal swab, and nasopharyngeal swab samples whose RNA was extracted via RNA extraction kit using primers for the detection of the *Orf1ab* gene of SARS-CoV-2**

During the second stage of this study, 71 patients with SARS symptoms had nasopharyngeal swabs, oropharyngeal swabs, saliva, and mouthwash with saline solution samples collected. Our results showed that among the four body biofluids collected, the nasopharyngeal swab was the only one which CT was maintained at a detectable range, and nasopharyngeal swabs had the highest viral load (**Figure 3E and 3F**). Diagnostic predictive values indicated that only nasopharyngeal swabs extracted using the RNA extraction kit can be used for the diagnosis of SARS-CoV-2 (**Supplementary Tables 2, 3, 4 and 5**).

### Diagnosis of SARS-CoV-2 via RT-LAMP after the extraction of sample RNA

We validated the newly developed RT-LAMP SARS-CoV-2 detection kit (Actin158, Orf9, and N5) by submitting 138 extracted RNA from nasopharyngeal swab samples for RT-LAMP diagnosis (**Figure 4A, 4B**, and **4C**). As expected, the internal control, Actin158, showed a positive result for all samples, except for the negative control (**Figure 4A**). Results obtained by RT-qPCR and RT-LAMP diagnosis is shown in **Supplementary Table 6**.

**Figure 4.**
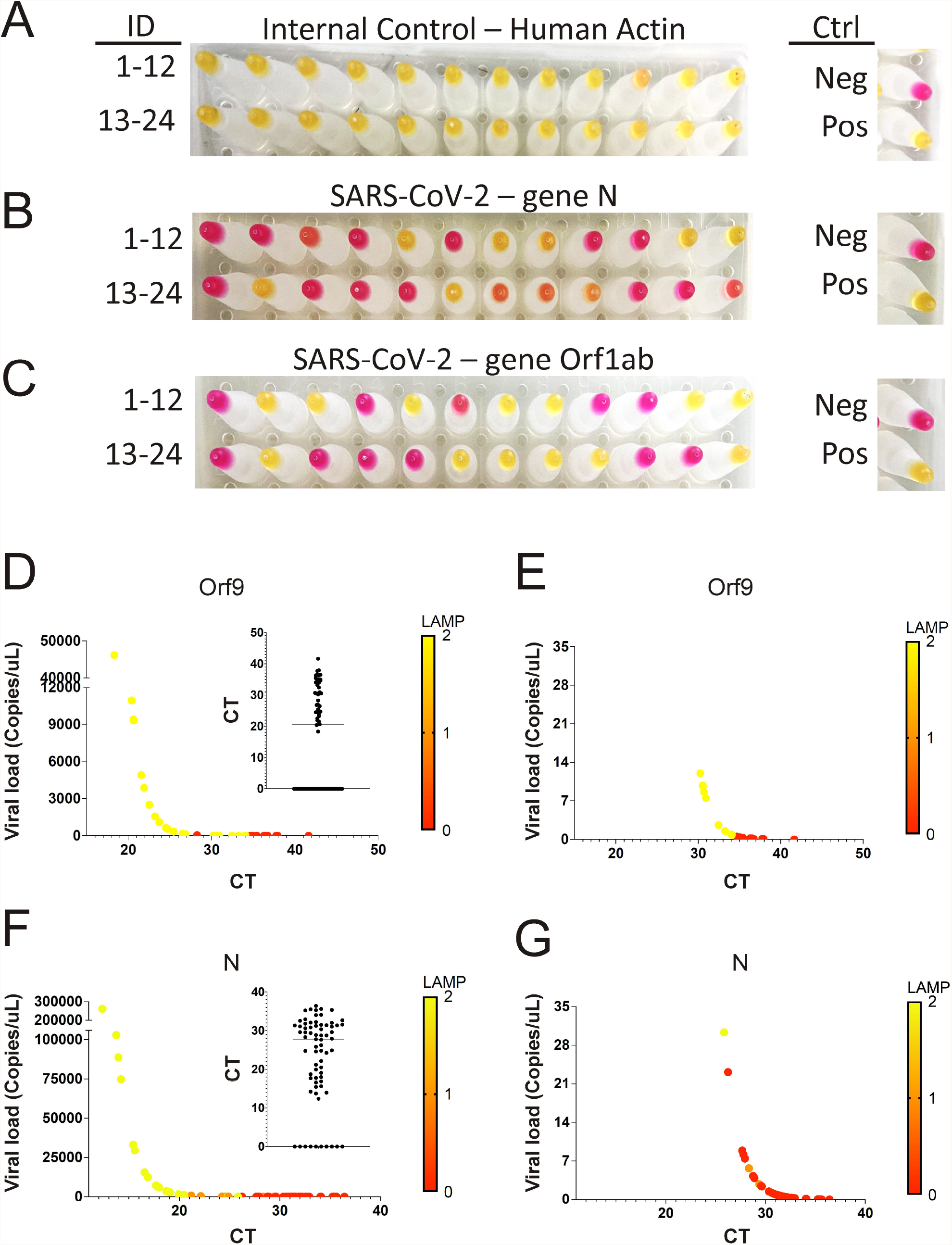
**Representative results of SARS-CoV-2 diagnosis via RT-qPCR and RT-LAMP from 71 human RNA samples extracted from nasopharyngeal swabs via an automated magnetic bead extraction kit. A-C - Representative RT-LAMP results from 24 human nasopharyngeal swabs whose RNA samples were extracted via automated magnetic bead extraction kit. D - RT-qPCR CT values and viral load using *Orf1ab* SARS-CoV-2 primers plotted against RT-LAMP diagnostic results using Orf9 primer set of nasopharyngeal swabs samples whose RNA samples were extracted via automated magnetic bead extraction kit. D’ - range of RT-qPCR CT values with median of all 71 nasopharyngeal swab. E - RT-qPCR CT values and viral load using *Orf1ab* SARS-CoV-2 primers plotted against RT-LAMP diagnostic results using Orf9 primer set of nasopharyngeal swabs samples whose RNA samples were extracted via automated magnetic bead extraction kit; the X axis indicates the maximum CT value of 35 for detection limit evaluation. F - RT-qPCR CT values and viral load using *N* SARS-CoV-2 primers plotted against RT-LAMP diagnostic results using N5 primer set of nasopharyngeal swabs samples whose RNA samples were extracted via automated magnetic bead extraction kit. F’ - range of RT-qPCR CT values with median of all 71 nasopharyngeal swab. G - RT-qPCR CT values and viral load using *N* SARS-CoV-2 primers plotted against RT-LAMP diagnostic results using N5 primer set of nasopharyngeal swabs samples whose RNA samples were extracted via automated magnetic bead extraction kit; the X axis indicates the maximum CT value of 35 for detection limit evaluation. ** RT-LAMP results were given a number from 0 to 2 according to the color of the reaction results. The pink color reactions were assigned the number 0, the orange color reactions were assigned the number 1, and the yellow color reactions were assigned the number 2. Each number and color are shown on the X-axis on the right.**

We compared the LAMP results to qPCR in terms of RT-qPCR CT value, viral load, and LAMP assay color (**Figure 4D** and **4F**). We observed that samples with a viral load higher than 1 copies/µL of *Orf1ab* were positive for SARS-CoV-2 via RT-qPCR assay using the primer set Orf9 (**Figure 4E**). Samples with a viral load higher than 28 copies/µL were positive for SARS-CoV-2 via the RT-LAMP assay using primer set N5 (**Figure 4G**).

Next, we calculated the sensitivity, specificity, positive predicted values (PPV), and negative predicted values (NPV) for RT-LAMP diagnosis using primer sets N5 and Orf9, comparing their diagnosis with the RT-qPCR diagnosis (Table 1). We considered all samples positive for viral load higher than 1 copies/µL, and the results are summarized in **Table 1**. Based on these results, we suggest that primer sets Orf9 and N5 should be used in combination for the diagnosis of SARS-CoV-2 in patients with SARS symptoms using nasopharyngeal swab samples. A representative diagnosis using combined analysis and results using the RT-LAMP SARS-CoV-2 detection kit is illustrated in **Supplementary Figure 1**. Sensitivity and specificity for combined diagnosis were 96% (95% CI 87-99) and 85% (95% CI 76-91), respectively. The PPV and NPV values for the combined diagnosis were 80% (95% CI 69-88) and 97% (95% CI 90-99), respectively.

**Table 1.** Predictive validity of the RT-LAMP SARS-CoV-2 detection kit in nasopharyngeal swab samples extracted by extraction kit.

| Extraction Kit - Nasopharyngeal Swab |  |  |  |  |  |  |  |
| --- | --- | --- | --- | --- | --- | --- | --- |
| RT-LAMP N5 |  |  |  |  |  |  |  |
| Sensitivity |  | Specificity |  | PPV |  | NPV |  |
| Value | 95% CI | Value | 95% CI | Value | 95% CI | Value | 95% CI |
| 0.78 | 0.65 to 0.87 | 0.95 | 0.88 to 0.98 | 0.91 | 0.79 to 0.96 | 0.88 | 0.80 to 0.93 |
| RT-LAMP Orf9 |  |  |  |  |  |  |  |
| Sensitivity |  | Specificity |  | PPV |  | NPV |  |
| Value | 95% CI | Value | 95% CI | Value | 95% CI | Value | 95% CI |
| 0.96 | 0.87 to 0.99 | 0.87 | 0.78 to 0.92 | 0.82 | 0.70 to 0.89 | 0.97 | 0.90 to 0.99 |
| RT-LAMP Orf9 and N5 |  |  |  |  |  |  |  |
| Sensitivity |  | Specificity |  | PPV |  | NPV |  |
| Value | 95% CI | Value | 95% CI | Value | 95% CI | Value | 95% CI |
| 0.96 | 0.87 to 0.99 | 0.85 | 0.76 to 0.91 | 0.8 | 0.69 to 0.88 | 0.97 | 0.90 to 0.99 |

## Discussion

In the State of Ceará, the official laboratory (LACEN-CE) routinely identifies the presence of SARS-CoV-2 in nasopharyngeal swabs via RT-qPCR after RNA extraction using commercial kits. On average, approximately 4 h are required for RNA extraction and qPCR test execution. However, it takes 48-72 hours to release official results. This time-consuming issue motivated us to pursue a new faster NAAT using RT-LAMP. This technique uses the Bst DNA polymerase, isolated in 1968 [20] from the thermophilic bacterium *Geobacillus stearothermophilus*, which has a strand displacement activity and requires a set of six primers.

The SARS-CoV-2 RT-LAMP NAAT developed here consists of three sets of primers, two of which are used for the identification of SARS-CoV-2 (N5 and Orf9), while one acts as the internal control to identify the human gene *actin* (Actin 158). The RT-LAMP reaction using viral samples from different SARS-CoV-2 variants showed that the N5 primer set had a lower limit of detection. Mutation rate analysis showed that the primer LB from the N5 set had a mutation rate of 16.8 %. *In silico* analyses showed that there is a guanine at P1 and GR clades, instead of a cytosine. A degenerate primer could recognize these variations improving the accuracy and efficiency of this NAAT in preliminary in vitro tests (**Supplementary figure 2**).

We tested this new NAAT on 138 nasopharyngeal swabs from SARS suspected patients whose RNA was extracted using a magnetic beads RNA extraction kit. Then, samples were submitted for diagnosis of SARS-CoV-2 using RT-qPCR and the new NAAT RT-LAMP developed for the diagnosis of SARS-CoV-2 using *N* and *Orf1ab* genes. The predictive diagnostic values obtained for PPV were greater than 80%, whereas the NPV values were greater than 85% for Orf9 and N5. Considering these results, we recommend using these sets of primers in combination for the diagnosis of SARS-CoV-2.

We also investigated the possibility of using an alternative methodology to simplify RNA extraction from patient samples. We tested the diagnosis of SARS-CoV-2 from nasopharyngeal swab samples using RNA extracted only by heat. However, we were unable to efficiently detect SARS-CoV-2 in these samples, most likely because the viral RNA content was not preserved after RNA extraction procedures. Considering this, we would not recommend using RNA extracted by the boiling method as template for the NAAT RT-LAMP developed here. Our study also identified that samples preserved in viral transport medium (VTM) interfered with the LAMP enzyme amplification reaction. Samples preserved in the VTM had a high rate of false positives. Thus, all samples were collected in PBS.

During the second stage of this study, samples of nasopharyngeal swabs, oropharyngeal swabs, saliva, and mouthwash in saline solution from SARS suspected patients were collected to test the predictive power of the new NAAT developed here. However, the results confirmed that only nasopharyngeal swab samples were suitable for the diagnosis of SARS-CoV-2 using the primer sets N5 and Orf9.

This study developed and validated a new nucleic acid amplification test for the diagnosis of SARS-CoV-2 using the RT-LAMP technique. The Orf9 and N5 primer sets showed low mutation rates, except for the primer LB from N5 set. Despite that, when evaluated in combination with the Orf9 primer set, our NAAT was able to maintain high sensitivity and specificity. This methodology could reduce the time required to release the official diagnosis by half as this new methodology does not require analysis of results using specific software programs.

## Data Availability

All data produced in the present study are available upon reasonable request to the authors

## Abbreviations

COVID-19: coronavirus disease 2019
FBS: fetal bovine serum
HMPV: human metapneumovirus
NAAT: nucleic acid amplification test
RT-LAMP: reverse transcription loop-mediated isothermal amplification
PBS: phosphate-buffered saline
RT-qPCR: quantitative reverse transcription-polymerase chain reaction
RV-16: rhinovirus serotype 16
SARS-CoV-2: severe acute respiratory syndrome-coronavirus 2
TCID_50_: median tissue culture infective dose
TPCK: tosylsulfonyl phenylalanyl chloromethyl ketone

## Author Contributions

Marco Clementino – Conceptualization, Data curation, Formal Analysis, Investigation, Methodology, Project Administration, Manuscript writing original draft, review and editing, Funding Acquisition.

Karene Ferreira Cavalcante, Vania Angelica Feitosa Viana, Dayara de Oliveira Silva, Caroline Rebouças Damasceno, Jessica Fernandes de Souza – Formal Analysis, Methodology, Validation, data curation. Rafhaella Nogueira Della Guardia Gondim, Daniel Macedo de Melo Jorge, Lyvia Maria Vasconcelos Carneiro Magalhães, Liana Perdigão Mello – Methodology, Project administration

Érico Antônio Gomes de Arruda, Roberto da Justa Pires Neto, Melissa Soares Medeiros – Medical assistance, investigation.

Armênio Aguiar dos Santos, Pedro Jorge Caldas Magalhães, Eurico Arruda, Aldo Ângelo Moreira Lima – Conceptualization, Project supervision, Investigation, Manuscript review and editing.

Alexandre Havt - Conceptualization, Data curation, Formal Analysis, Investigation, Methodology, Project Administration, Manuscript writing original draft, review and editing, Funding Acquisition

