## Supplemental files for "Detection of SARS-CoV-2 in Different Human Biofluids Using the Loop-Mediated Isothermal Amplification Assay: A Prospective Diagnostic Study in Fortaleza, Brazil"

**Supplementary table 1. Demographics of patients.** *
* Percentage may not totalize 100 because of rounding. ** IQR denotes interquartile range.

| Population of Stage 1 | Number of patients N = 71 |
|---|---|
| **Age in years – median (IQR) ** ** | 37 (29) |
| **Days of symptoms – Average (STDEV)** | 4 (3) |
| **Male sex – no. (%)** | 34/70 (49) |
| **City of Residency in the state of Ceará – no. / total (%)** | |
| 1.Fortaleza | 67 / 70 (96) |
| 1.Caucaia | 1 / 70 (1) |
| 1.Maracanaú | 1 / 70 (1) |
| 1.Maranguape | 1 / 70 (1) |
| **Health care unit – no. / total no. (%)** | |
| 1.Barra do Ceará | 19 / 70 (27) |
| 1.José Walter | 17 / 70 (18) |
| 1.Hospital da Criança | 15 / 70 (21) |
| 1.Barra do Futuro | 5 / 70 (7) |
| 1.Cristo Redentor | 5 / 70 (7) |
| 1.Others | 9 / 70 (13) |

| Population of Stage 2 | Number of patients N = 71 |
|---|---|
| **Age in years – median (IQR) ** ** | 39 (27) |
| **Days of symptoms – Average (STDEV)** | 5 (2) |
| **Male sex – no. (%)** | 39/71 (55) |
| **City of Residency in the state of Ceará – no. / total (%)** | |
| 1. Fortaleza | 63 / 71 (89) |
| 2. Maranguape | 4/ 71 (6) |
| 3. Maracanaú | 2 / 71 (3) |
| 4. Iguatu | 1 / 71 (1) |
| **Health care unit – no. / total no. (%)** | |
| 1.Hospital São José | 71 / 71 (100) |

### Supplementary results table 2– RT-LAMP SARS-CoV-2 detection kit results in human biofluid samples

**Oropharyngeal swab**

| LAMP - N5 | Viral load (Copies/uL) by RT-qPCR (N=70) | | | Diagnostics RT-LAMP (N=70) | | | | | |
|---|---|---|---|---|---|---|---|---|---|
| CT range | Samples | Average | STDEV | Positive | Negative | TP | FP | TN | FN |
| 1 - 20 | 0.0 | 0.0 | 0.0 | 0.0 | 0.0 | 0.0 | 0.0 | 0.0 | 0.0 |
| 20 - 30 | 0.0 | 0.0 | 0.0 | 0.0 | 0.0 | 0.0 | 0.0 | 0.0 | 0.0 |
| 30 - 40 | 27.0 | 1.3 | 2.0 | 5.0 | 22.0 | 4.0 | 1.0 | 7.0 | 15.0 |
| ND | 43.0 | 0.0 | 0.0 | 3.0 | 40.0 | 0.0 | 3.0 | 34.0 | 6.0 |

| LAMP - Orf9 | Viral load (Copies/uL) by RT-qPCR (N=70) | | | Diagnostics RT-LAMP (N=70) | | | | | |
|---|---|---|---|---|---|---|---|---|---|
| CT range | Patients | Average | STDEV | Positive | Negative | TP | FP | TN | FN |
| 1 - 20 | 0.0 | 0.0 | 0.0 | 0.0 | 0.0 | 0.0 | 0.0 | 0.0 | 0.0 |
| 20 - 30 | 0.0 | 0.0 | 0.0 | 0.0 | 0.0 | 0.0 | 0.0 | 0.0 | 0.0 |
| 30 - 40 | 27.0 | 1.3 | 2.0 | 7.0 | 20.0 | 5.0 | 2.0 | 6.0 | 14.0 |
| ND | 43.0 | 0.0 | 0.0 | 3.0 | 40.0 | 0.0 | 3.0 | 34.0 | 6.0 |

**Saliva**

| LAMP - N5 | Viral load (Copies/uL) by RT-qPCR (N=69) | | | Diagnóstics RT-LAMP (N=69) | | | | | |
|---|---|---|---|---|---|---|---|---|---|
| CT range | Patients | Average | STDEV | Positive | Negative | TP | FP | TN | FN |
| 1 - 20 | 0.0 | 0.0 | 0.0 | 0.0 | 0.0 | 0.0 | 0.0 | 0.0 | 0.0 |
| 20 - 30 | 12.0 | 441.7 | 753.0 | 11.0 | 1.0 | 10.0 | 0.0 | 0.0 | 1.0 |
| 30 - 40 | 23.0 | 3.4 | 4.5 | 5.0 | 18.0 | 3.0 | 2.0 | 11.0 | 7.0 |
| ND | 34.0 | 0.0 | 0.0 | 1.0 | 32.0 | 0.0 | 1.0 | 30.0 | 3.0 |

| LAMP - Orf9 | Viral load (Copies/uL) by RT-qPCR (N=69) | | | Diagnostics RT-LAMP (N=69) | | | | | |
|---|---|---|---|---|---|---|---|---|---|
| CT range | Patients | Average | STDEV | Positive | Negative | TP | FP | TN | FN |
| 1 - 20 | 0.0 | 0.0 | 0.0 | 0.0 | 0.0 | 0.0 | 0.0 | 0.0 | 0.0 |
| 20 - 30 | 12.0 | 441.7 | 753.0 | 7.0 | 5.0 | 7.0 | 0.0 | 0.0 | 5.0 |
| 30 - 40 | 23.0 | 3.4 | 4.5 | 8.0 | 15.0 | 4.0 | 3.0 | 9.0 | 7.0 |
| ND | 33.0 | 0.0 | 0.0 | 6.0 | 27.0 | 0.0 | 5.0 | 24.0 | 4.0 |

**Saline wash**

| LAMP - N5 | Viral load (Copies/uL) by RT-qPCR (N=68) | | | Diagnostics RT-LAMP (N=68) | | | | | |
|---|---|---|---|---|---|---|---|---|---|
| CT range | Patients | Average | STDEV | Positive | Negative | TP | FP | TN | FN |
| 1 - 20 | 0.0 | 0.0 | 0.0 | 0 | 0 | 0 | 0 | 0 | 0 |
| 20 - 30 | 3.0 | 1562.2 | 2174.3 | 3 | 0 | 3 | 0 | 0 | 0 |
| 30 - 40 | 23.0 | 3.4 | 3.6 | 8 | 15 | 7 | 1 | 7 | 8 |
| ND | 42.0 | 0.0 | 0.0 | 7 | 35 | 2 | 5 | 30 | 5 |

| LAMP - Orf9 | Viral load (Copies/uL) by RT-qPCR (N=68) | | | Diagnostics RT-LAMP (N=68) | | | | | |
|---|---|---|---|---|---|---|---|---|---|
| CT range | Patients | Average | STDEV | Positive | Negative | TP | FP | TN | FN |
| 1 - 20 | 0.0 | 0.0 | 0.0 | 0 | 0 | 0 | 0 | 0 | 0 |
| 20 - 30 | 3.0 | 1562.2 | 2174.3 | 3 | 1 | 2 | 0 | 0 | 1 |
| 30 - 40 | 23.0 | 3.4 | 3.6 | 12 | 11 | 10 | 2 | 6 | 5 |
| ND | 42.0 | 0.0 | 0.0 | 3 | 39 | 2 | 1 | 36 | 3 |

#### Supplementary table 3 – Predictive validity of the RT-LAMP SARS-CoV-2 detection kit in human biofluid samples

| Oropharyngeal swab | | | | | | | |
|---|---|---|---|---|---|---|---|
| **RT-LAMP Orf9** | | | | | | | |
| Sensitivity | | Specificity | | PPV | | NPV | |
| Value | 95% CI | Value | 95% CI | Value | 95% CI | Value | 95% CI |
| 0.2 | 0.08 to 0.39 | 0.88 | 0.76 to 0.95 | 0.5 | 0.23 to 0.76 | 0.66 | 0.54 to 0.77 |
| **RT-LAMP N5** | | | | | | | |
| Sensitivity | | Specificity | | PPV | | NPV | |
| Value | 95% CI | Value | 95% CI | Value | 95% CI | Value | 95% CI |
| 0.16 | 0.06 to 0.34 | 0.91 | 0.79 to 0.96 | 0.5 | 0.21 to 0.78 | 0.66 | 0.53 to 0.76 |

| Saliva | | | | | | | |
|---|---|---|---|---|---|---|---|
| **RT-LAMP Orf9** | | | | | | | |
| Sensitivity | | Specificity | | PPV | | NPV | |
| Value | 95% CI | Value | 95% CI | Value | 95% CI | Value | 95% CI |
| 0.4 | 0.24 to 0.59 | 0.8 | 0.65 to 0.89 | 0.57 | 0.36 to 0.76 | 0.67 | 0.53 to 0.78 |
| **RT-LAMP - N5** | | | | | | | |
| Sensitivity | | Specificity | | PPV | | NPV | |
| Value | 95% CI | Value | 95% CI | Value | 95% CI | Value | 95% CI |
| ND | 0.35 to 0.72 | 0.93 | 0.81 to 0.97 | 0.81 | 0.56 to 0.93 | 0.78 | 0.65 to 0.87 |

| Saline wash | | | | | | | |
|---|---|---|---|---|---|---|---|
| RT-LAMP Orf9 | | | | | | | |
| Sensitivity | | Specificity | | PPV | | NPV | |
| Value | 95% CI | Value | 95% CI | Value | 95% CI | Value | 95% CI |
| 0.6 | 0.4 to 0.77 | 0.93 | 0.82 to 0.97 | 0.82 | 0.58 to 0.93 | 0.82 | 0.69 to 0.90 |
| RT-LAMP - N5 | | | | | | | |
| Sensitivity | | Specificity | | PPV | | NPV | |
| Value | 95% CI | Value | 95% CI | Value | 95% CI | Value | 95% CI |
| 0 | 0.3 to 0.66 | 0 | 0.72 to 0.93 | 0 | 0.43 to 0.83 | 0.74 | 0.60 to 0.84 |

#### Supplementary results table 4 – RT-LAMP SARS-CoV-2 detection kit results in nasopharyngeal swab samples extracted by heat

| Data from Samples Extracted by Heat | | | | | | | | | |
|---|---|---|---|---|---|---|---|---|---|
| Nasopharyngeal Swab | | | | | | | | | |
| LAMP - N5 | Viral load (Copies/uL) by RT-qPCR (N=71) | | | Diagnostics RT-LAMP (N=71) | | | | | |
| CT range | Samples | Average | STDEV | Positive | Negative | TP | FP | TN | FN |
| 1 - 20 | 1 | 8247.8 | 0.0 | 1 | 0 | 0 | 1 | 0 | 0 |
| 20 - 30 | 8 | 809.1 | 1346.9 | 8 | 0 | 3 | 5 | 0 | 0 |
| 30 - 40 | 28 | 0.6 | 1.3 | 14 | 14 | 5 | 9 | 10 | 4 |
| ND | 34 | 0.0 | 0.0 | 8 | 26 | 2 | 6 | 14 | 12 |
| LAMP - Orf9 | Viral load (Copies/uL) by RT-qPCR (N=71) | | | Diagnostics RT-LAMP (N=71) | | | | | |
| CT range | Samples | Average | STDEV | Positive | Negative | TP | FP | TN | FN |
| 1 - 20 | 0 | 0.0 | 0.0 | 0 | 0 | 0 | 0 | 0 | 0 |
| 20 - 30 | 11 | 901.6 | 1832.5 | 10 | 11 | 3 | 7 | 0 | 1 |
| 30 - 40 | 24 | 2.4 | 3.7 | 17 | 7 | 6 | 11 | 4 | 3 |
| ND | 36 | 0.0 | 0.0 | 24 | 12 | 10 | 14 | 8 | 4 |

#### Supplementary table 5 – Predictive validity of the RT-LAMP SARS-CoV-2 detection kit in nasopharyngeal swab samples extracted by heat

| Nasopharyngeal swab extracted by heat | | | | | | | |
|---|---|---|---|---|---|---|---|
| RT-LAMP N5 | | | | | | | |
| Sensitivity | | Specificity | | PPV | | NPV | |
| Value | 95% CI | Value | 95% CI | Value | 95% CI | Value | 95% CI |
| 0.38 | 0.22 to 0.57 | 0.53 | 0.39 to 0.67 | 0.32 | 0.18 to 0.49 | 0.6 | 0.44 to 0.73 |
| RT-LAMP Orf9 | | | | | | | |
| Sensitivity | | Specificity | | PPV | | NPV | |
| Value | 95% CI | Value | 95% CI | Value | 95% CI | Value | 95% CI |
| 0.7 | 0.51 to 0.84 | 0.27 | 0.16 to 0.41 | 0.37 | 0.25 to 0.50 | 0.6 | 0.38 to 0.78 |

### Supplementary Table 6 – RT-LAMP SARS-CoV-2 detection kit results in nasopharyngeal swab samples extracted by extraction kit

| Data from Samples Extracted by Extraction Kit | | | | | | | | | |
|---|---|---|---|---|---|---|---|---|---|
| Nasopharyngeal Swab | | | | | | | | | |
| **LAMP - N5** | Viral load (Copies/uL) by RT-qPCR (N = 138) | | | Diagnostics RT-LAMP (N = 138) | | | | | |
| CT range | Samples | Average | STDEV | Positive | Negative | TP | FP | TN | FN |
| 1 - 20 | 3 | 31257.9 | 11315.7 | 2 | 1 | 2 | 0 | 0 | 1 |
| 20 - 30 | 30 | 2228.8 | 2971.0 | 30 | 0 | 30 | 0 | 0 | 0 |
| 30 - 40 | 40 | 2.2 | 3.3 | 11 | 29 | 9 | 2 | 19 | 10 |
| ND | 65 | 0.0 | 0.0 | 2 | 63 | 0 | 2 | 63 | 0 |
| **LAMP - Orf9** | Viral load (Copies/uL) by RT-qPCR (N = 138) | | | Diagnostics RT-LAMP (N = 138) | | | | | |
| CT range | Samples | Average | STDEV | Positive | Negative | TP | FP | TN | FN |
| 1 - 20 | 3 | 31257.9 | 11315.7 | 2 | 1 | 2 | 0 | 0 | 1 |
| 20 - 30 | 30 | 2228.8 | 2971.0 | 30 | 0 | 30 | 0 | 0 | 0 |
| 30 - 40 | 40 | 2.2 | 3.3 | 25 | 15 | 19 | 6 | 14 | 1 |
| ND | 65 | 0.0 | 0.0 | 5 | 60 | 0 | 5 | 60 | 0 |
| **LAMP - Orf9 and N5** | Viral load (Copies/uL) by RT-qPCR (N = 138) | | | Diagnostics RT-LAMP (N = 138) | | | | | |
| CT range | Samples | Average | STDEV | Positive | Negative | TP | FP | TN | FN |
| 1 - 20 | 3 | 31257.9 | 11315.7 | 2 | 1 | 2 | 0 | 0 | 1 |
| 20 - 30 | 30 | 2228.8 | 2971.0 | 30 | 0 | 30 | 0 | 0 | 0 |
| 30 - 40 | 40 | 2.2 | 3.3 | 24 | 26 | 19 | 6 | 14 | 1 |
| ND | 65 | 0.0 | 0.0 | 6 | 59 | 0 | 6 | 59 | 0 |

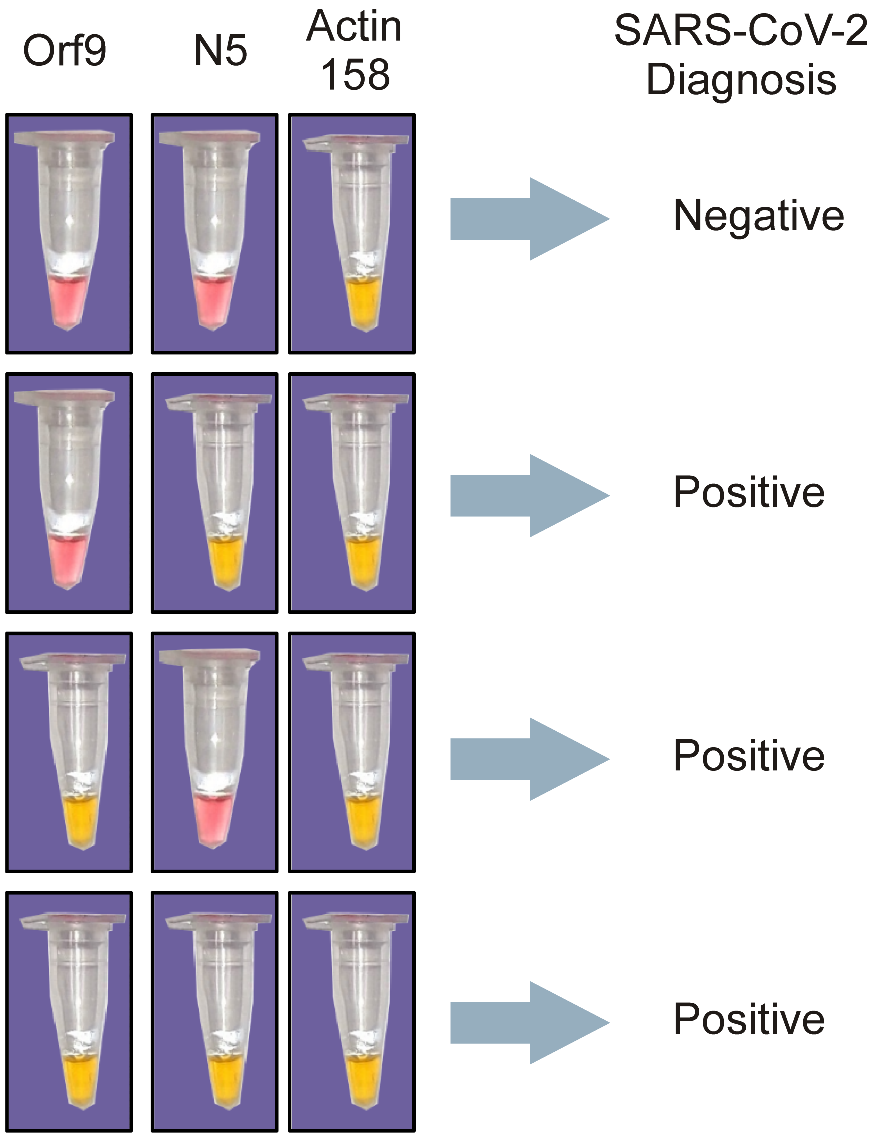

**Supplementary figure 1. Representative diagnostic results for detection of SARS-CoV-2 using the RT-LAMP kit developed.**

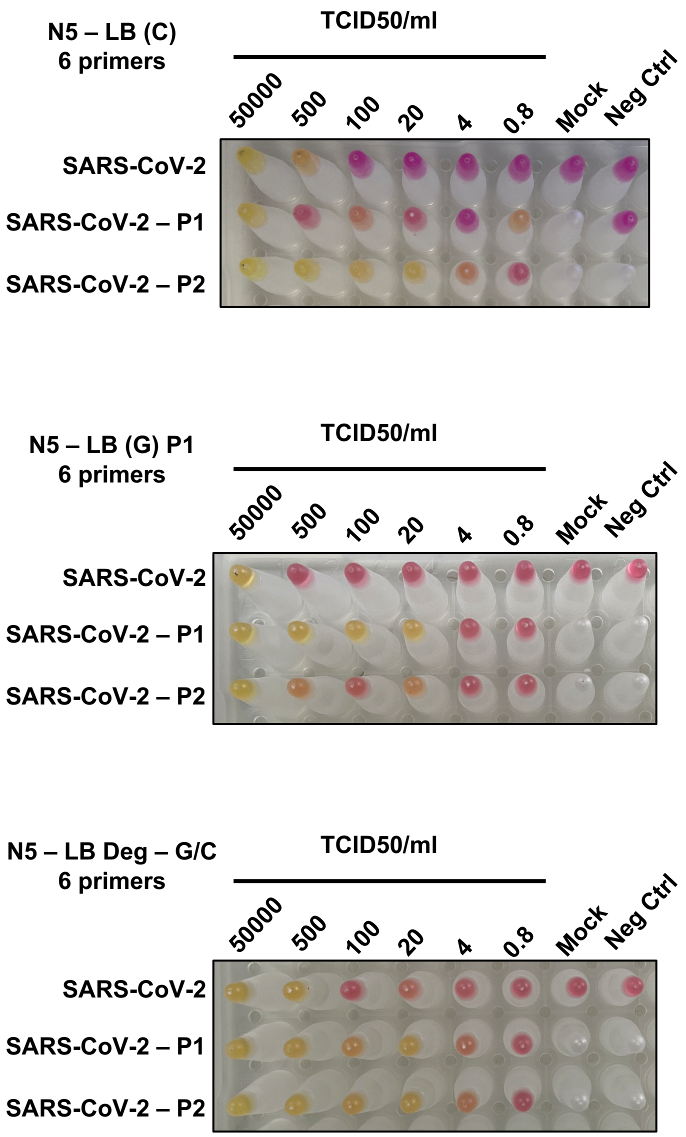

**Supplementary figure 2.** Representative limit of detection results of the RT-LAMP reaction using the N5 primer set in samples of SARS-CoV-2, Variant P1 and variant P2 cultured in vitro.
